# Epidemiological Study of COVID-19 Infections: Case of Ga East Municipal Hospital Treatment Centre - Kwabenya-Ghana

**DOI:** 10.1101/2021.08.11.21261917

**Authors:** Ebenezer Oduro-Mensah, Delali Asubonteng, Shirley Crankson

## Abstract

**Background:** The COVID-19 pandemic still poses a considerable threat to global health, resulting in an unprecedented demand for regular research to continuously identify and update its risk profiles to ensure relevant interventions.

**Objectives:** This study aimed to examine the epidemiological characteristics of the COVID-19 outbreak in Ghana and identify risk factors for severe COVID-19 disease.

**Method:** This was a cross-sectional study involving the data of patients with COVID-19 clinically managed at the Ga-East Municipal Hospital, the main COVID-19 treatment centre in Ghana, from 21st March to 21st June 2020. The data were retrieved from the electronic medical records and folders of the patients. It included sociodemographic characteristics, COVID-19 disease severity, and treatment outcomes. Binomial logistic regression was used to identify risk factors of severe COVID-19 illness.

**Result:** Among the 360 COVID-19 cases in this study, 55.3% were males, and 44.7% were females. Their mean age was 39.9±16.7years. Most of them were Ghanaians (92.8%) and employed (72.5%). The majority (93%) presented with mild disease, and hypertension (19.2%) was the most common comorbidity. The average length of hospital admission was 10.6±6.4day. Many of the cases recovered (98.6%), resulting in a case fatality of 1.4%. Finally, the logistic regression showed that increasing age (OR=1.12, p= 0.002) and diabetes mellitus (OR=19.85, p=0.007) are risk factors for severe COVID-19 disease.

**Conclusion:** Findings from the study confirmed that increasing age and diabetes mellitus are risk factors for severe COVID-19 disease. Thus, Ghana could prioritise these identified populations when implementing interventions to reduce the COVID-19 disease burden.

**Strength and limitations of this study:**

- This study is one of the few studies exploring the risk factors for severe COVID-19 disease in Ghana; thus, it presents novel context-specific findings that could inform COVID-19 case management in Ghana.
- The study was limited in drawing a causal association between the explanatory and outcome variables due to the cross-sectional design.
- The study included only one COVID-19 treatment centre. Consequently, the findings may not be generalizable to the entire COVID-19 population in Ghana, even though the included centre is the main COVID-19 treatment centre.

## Introduction

COVID-19 cases keep increasing since its discovery in December 2019. The disease which first broke out in Wuhan, China, is caused by a pathogen that primarily targets the human respiratory system. The world health organisation (WHO) declared its outbreak as an international public health emergency on 30th January 2020. Since its emergence, more than two hundred million cases and over four million deaths have been confirmed globally [1]. Ghana recorded its first two cases, which were imported, on 12th March 2020, a day after WHO declared the outbreak a pandemic. Even though the country had the advantage of retrospection, by observing the dynamics of the pandemic from countries that had earlier cases, the outbreak still had a mammoth impact on the health and economy of the country. As of 31st July 2021, Ghana had confirmed nearly one hundred and thousand cases and about eight hundred deaths [2].

After recording its first cases, the Ghana government, through the Ghana Health Service (GHS), implemented several measures to control and avert any further spread of the COVID-19 virus in Ghana. These measures included lockdowns in the country’s COVID-19 disease hotspots, increased testing capacities of earmarked COVID-19 and revamped local enterprises to produce affordable nose masks. Others were rigorous contact tracing, identification of isolation centres, incentivisation of front-line workers, intensive public education on the attributes of the COVID-19 virus, personal hygiene, and the influence of crowding on the viral spread (2).

Even though these measures were lauded as efficient interventions, some of them, precisely the lockdown, were thought to be drastic (20). This was because it exposed the country to the other looming threat of the outbreak, crippling personal and institutional finances. The country’s fragile economy and diverse socioeconomic characteristics made this imminent consequence very daunting. However, the words of the country’s president: ‘We know how to bring the economy back to life, but We do not know how to bring people back to life’ justified the application of the lockdown. Nonetheless, the impact of these measures is yet to be empirically assessed to determine their continued relevance as the pandemic rages.

Following the COVID-19 outbreak, several studies have reported country/population-specific risk characteristics of the pandemic to inform setting suitable interventions. Most of these regular context-based studies informed the critical shielding interventions implemented at the initial and subsequent stages of the outbreak in the specific populations. Thus, it is prudent that Ghana also reports on the dynamics of the COVID-19 pandemic to inform and update current interventions to alleviate the disease burden. Therefore, this study aimed to explore the COVID-19 disease burden in Ghana, particularly its epidemiological characteristics, to inform context-based interventions, as done in other jurisdictions. Additionally, the study aimed to address the research dearth in Ghana’s COVID-19 literature space by profiling the risk factors associated with COVID-19 disease severity.

## Methods

### Data source

This cross-sectional study was conducted at Ga-East Municipal Hospital (GEMH), the main centre earmarked for COVID-19 treatment in Ghana. Data was accessed from the folder and electronic medical records of all patients admitted at GEMH from 21st March to 21st June with Polymerase Chain Reaction (PCR)-confirmed COVID-19 diagnosis. Consequently, patients admitted at GEMH with no PCR-confirmed COVID-19 diagnosis were excluded from this study. The data were collected using a data collection template developed a priori by the researchers. The collected data included age, gender, comorbidities, disease severity classification, and socioeconomic status. The study sample size was 360.

### Variables

#### Outcome Variables

Two outcomes were analysed in this study: COVID-19 disease classification (CDC), the primary study outcome, and COVID-19 treatment outcome (CTO). The CDC variable described the severest disease classification of the patient during the illness experience. This study classified this variable into four main categories based on the WHO guidelines on COVID-19 disease classification (4). The categories were mild, moderate, severe, and critical diseases.

Those classified as having a mild disease were symptomatic patients meeting the case definition for COVID-19 (low-grade fever, dry cough, sore throat, nasal congestion, headache, myalgia, malaise) without evidence of viral pneumonia or hypoxia. Those classified as moderate were adolescents or adults with clinical signs of pneumonia (fever, cough, dyspnoea, fast breathing) but no symptoms of severe pneumonia, including SpO2 ≥ 94% on room air (SpO2 level chosen for Ghana). It also included children with non-severe pneumonia but have a cough or difficulty and fast breathing. Patients with moderate symptoms usually required no supplemental oxygen.

Severe diseases included adolescent or adult with clinical signs of pneumonia (fever, cough, dyspnoea, fast breathing) plus one of the following: respiratory rate > 30 breaths/min; severe respiratory distress; or SpO2 < 94% on room air. Children with a cough or difficulty breathing, plus at least one of the following, central cyanosis or SpO2 < 90% and severe respiratory distress, were also classified to have severe COVID-19 disease. Patients with severe COVID-19 disease usually require supplemental oxygen support in a high dependency unit.

Those classified as having critical COVID-19 disease were patients developing Acute Respiratory Distress Syndrome (ARDS), sepsis, septic shock, acute thrombosis, or acute kidney injury after laboratory confirmation of COVID-19 infection. Such patients may require intubation and ventilation to maintain life over the illness period.

Therefore, this study described the characteristics of the study population based on these four CDC in the descriptive analysis. However, based on the overarching aim of this study, the mild and moderate groups were merged to represent non-severe COVID-19 disease, and the severe and critical groups were combined to represent severe COVID-19 disease in further analysis. Thus, this study explored the CDC variable as a binary outcome in the subsequent analysis. Therefore, it was coded as 0 – ‘non-severe COVID-19 disease’ (mild/moderate symptoms) and 1 – ‘severe COVID-19 disease’ (severe/critical symptoms).

The CTO variable described whether the patient died or recovered from the COVID-19 treatment. Therefore, it was also explored as a binary variable, with ‘recovered’ coded as 0 and ‘died’ 1.

#### Explanatory variables

Age, gender, comorbidities, education, employment, nationality, and marital status of the COVID-19 cases from GEMH were explored as independent variables in this study. These variables were based on evidence from current literature on potential risk factors of COVID-19 severity and mortality [5,6,7]. Age was calculated from the patient’s date of birth and assessed as a continuous variable, and gender was specified as either male or female. Comorbidity was described as whether the patient had other specific underlying chronic conditions, such as diabetes, hypertension, chronic obstructive pulmonary disease (COPD), tuberculosis, HIV/AIDs, cancer, and chronic steroids use or not. Employment status was defined as either employed or unemployed., and education level was categorised into four domains: No formal education, primary, secondary, and tertiary education. Marital status was also specified as either single, married, cohabiting, divorced/separated or widowed.

### Data analysis

Descriptive, bivariate, and multivariate analyses were conducted with SPSS software version The level of significance was set at p≤0.05 and 95% Confidence Interval (CI). Descriptive analysis was used to summarise the characteristics of the COVID-19 cases. Further, bivariate analysis was performed to examine the associations between each explanatory variable and the dependent variables. Chi-square, Spearman’s rho, and Mann-Whitney U test were used for the bivariate analysis. Finally, binomial logistic regression was conducted to examine risk factors for severe COVID-19 illness.

## Results

The sample had three hundred and sixty (360) cases, of which 55.3% were males, and 44.7% were females. Their ages ranged from 0 to 83 years, with a mean age of 39.9±16.7years. Most of them (92.8%) were Ghanaians, married (48.3%), employed (72.5%) and had received secondary/vocational training (42.5%). The top three comorbidities were hypertension (19.2%), hypertension and diabetes combined (7.2%) and pulmonary diseases (4.2%). With COVID-19 disease classification, about 93% of the cases had mild symptoms, and almost 4% had severe symptoms. Also, while no male had critical COVID-19 symptoms, more males (57%) had severe COVID-19 symptoms than females (43%).

Furthermore, most (29%) of the severe COVID-19 symptoms were among those with hypertension and diabetes. Averagely, 99% of the cases recovered, and 1% died. Among those who died, four had severe symptoms while one had critical COVID-19 symptoms (table 1). On hospital admission length, cases spent 0 to 38 days on admission, with an average hospital stay of 10.6±6.4days.

**Table 1:**
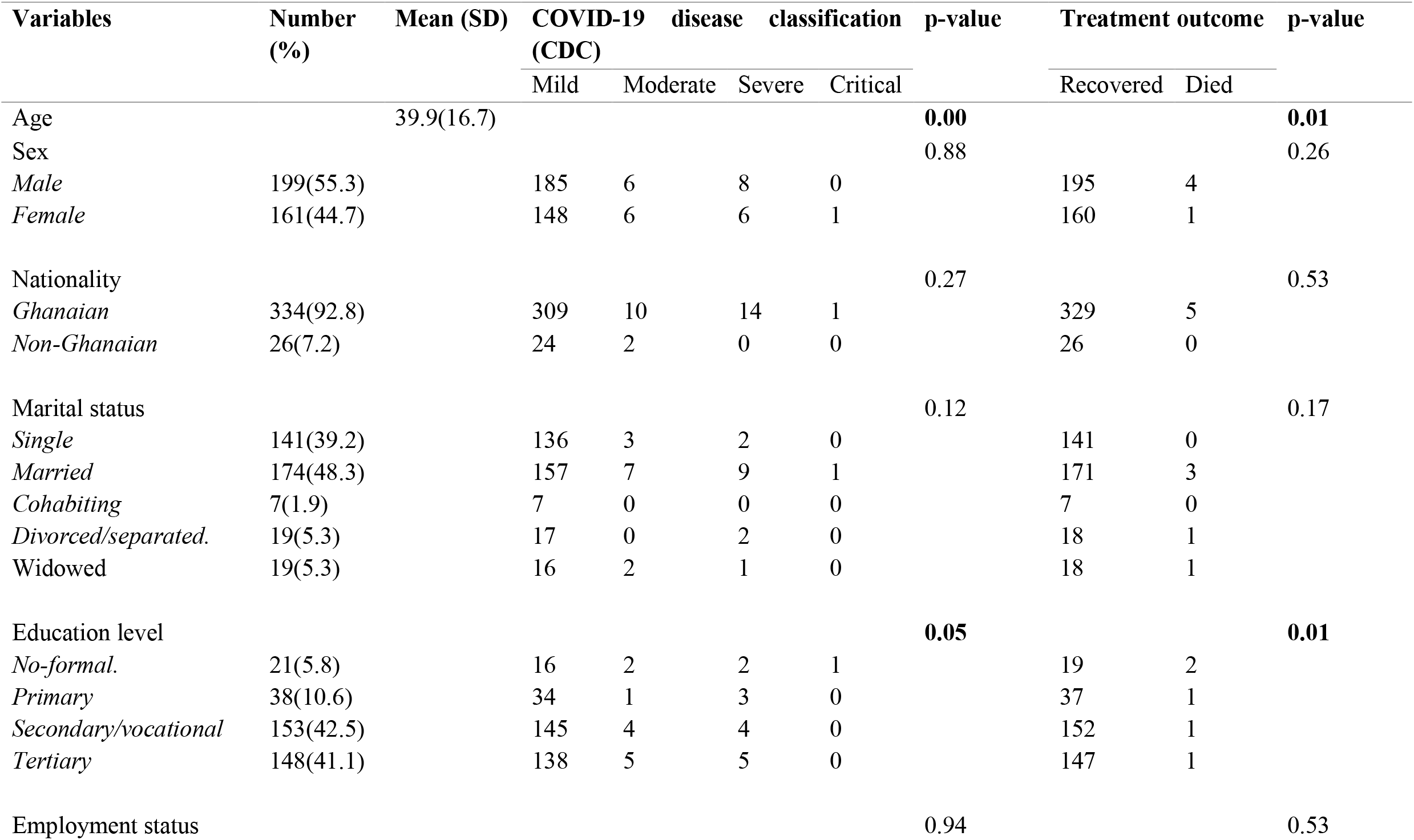
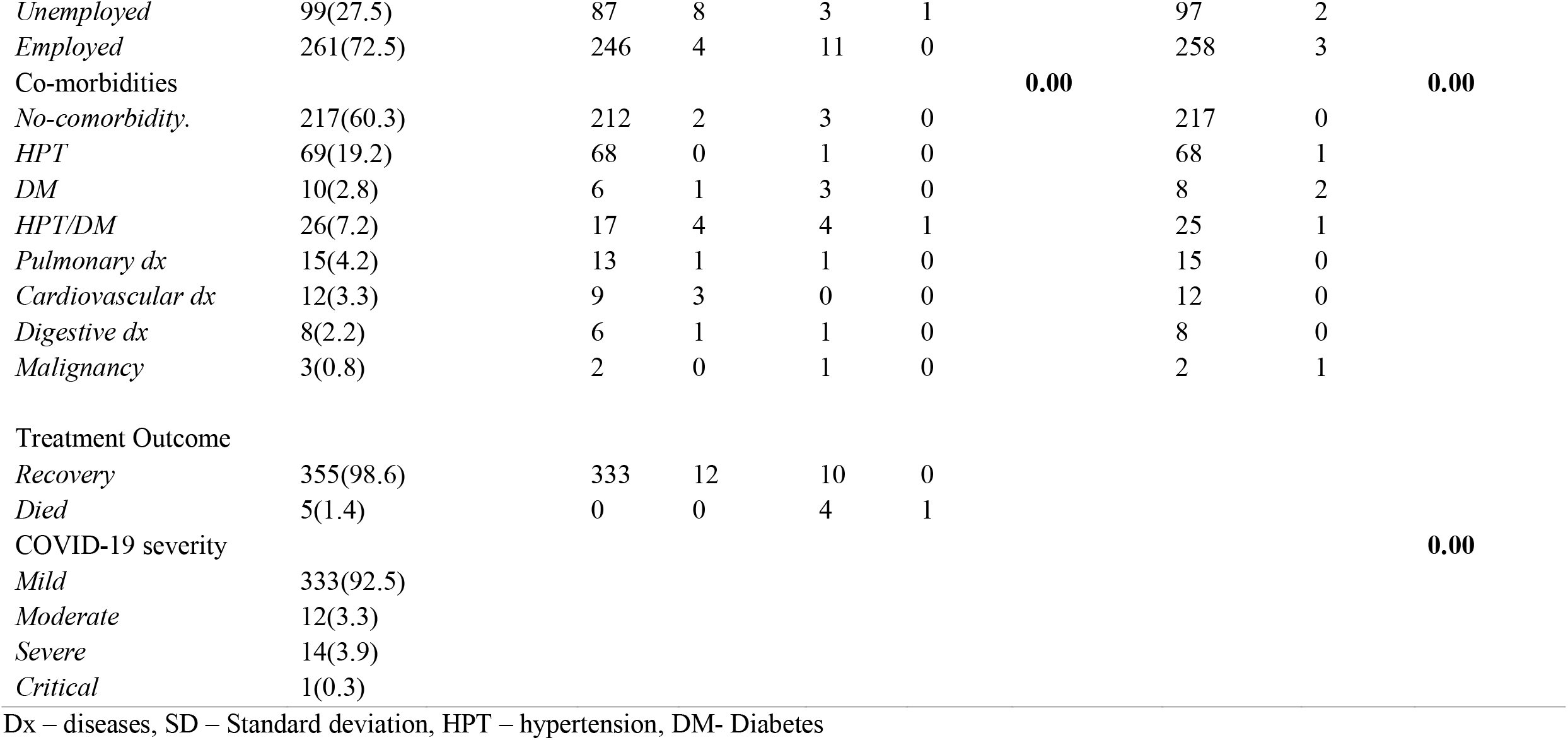
Sample characteristics and findings of bivariate analysis (N=360)

Findings of the bivariate analysis indicated that age (p=0.00), education level (0.05) and presence of comorbidities (p=0.00) are significantly associated with COVID-19 disease. Similarly, age (p=0.01), education level (0.01), comorbidities (p=0.00) and COVID-19 disease severity (p=0.00) are correlated with COVID-19 treatment outcomes (table 1). In the binomial logistic regression, increasing age (OR=1.12, p= 0.002) was identified to increase the likelihood of developing severe COVID-19 disease. Likewise, the presence of diabetes mellitus was associated with higher odds of severe COVID-19 disease (OR=19.85, p=0.007) – table 2.

**Table 2:**
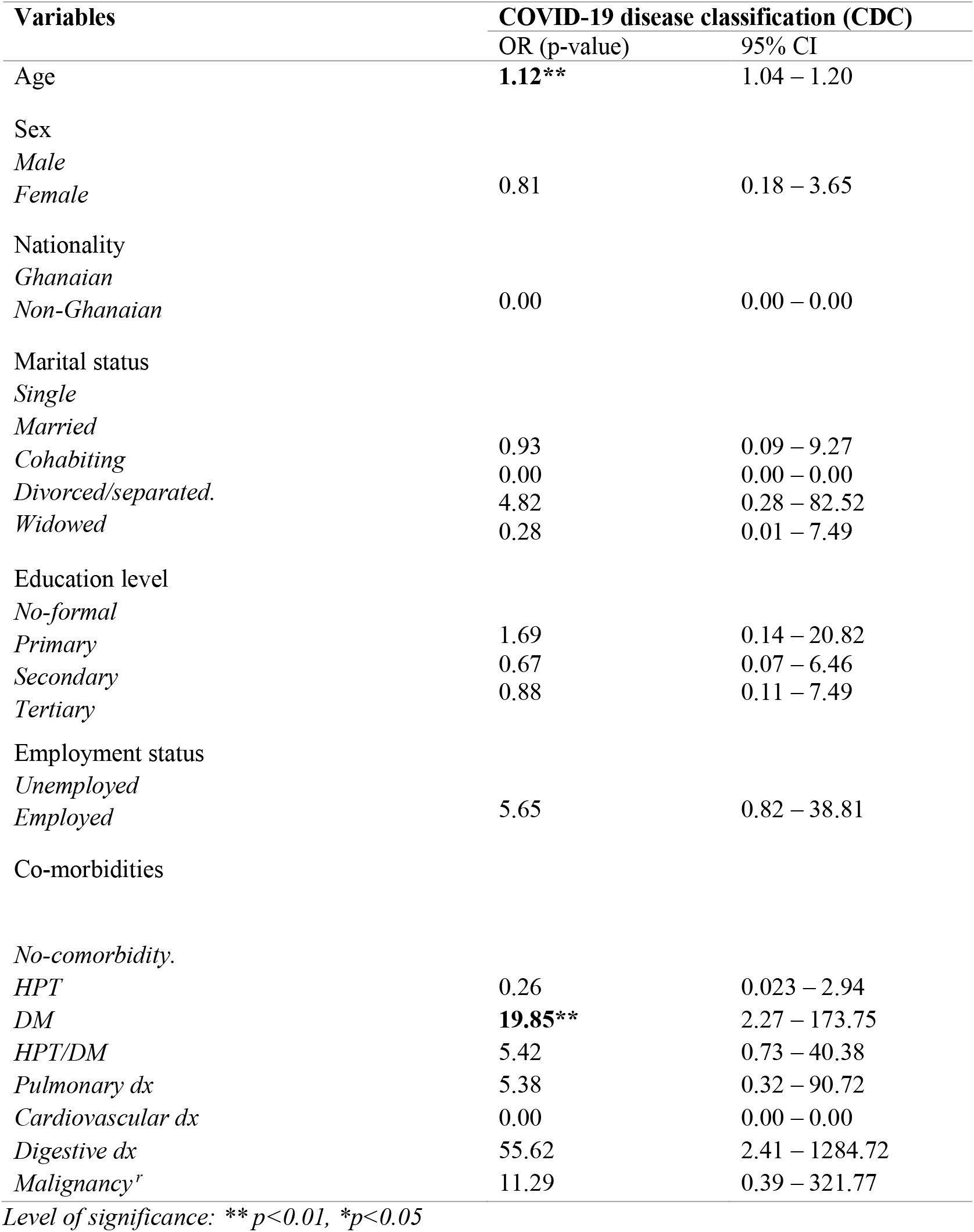
Binomial logistic regression of risk factors for COVID-19 disease

## Discussion

The study aimed to describe and explore the COVID-19 disease burden in Ghana. Data was sourced from the main COVID-19 treatment centre in Ghana - GEMH. As already indicated, GEMH started managing confirmed cases of COVID-19 from the 21st of March 2020, two weeks after Ghana confirmed its first two cases. The first two confirmed cases necessitated the need for government to scout for a treatment centre for the onslaught of the virus. Accordingly, GEMH was settled on for the management of subsequent cases. Before that, most of the confirmed cases were seen at the Greater Accra Regional Hospital, Tema General Hospital, and the homes of the patients. In terms of COVID-19 infection distribution, most of the first patients at Ghana were imported from other countries, including Norway and the UK [2]. Following the expected community spread, infection dynamics changed from imported to local cases resulting in the increased local COVID-19 cases being managed at GEMH.

Regarding the disease burden, the data from GEMH showed that Ghana recorded a relatively lower COVID-19 mortality rate (less than 2%) during the early stage of the virus outbreak. Similarly, less than 5% of the admitted cases experienced severe to critical symptoms associated with the disease. These findings may be related to the rigorous actions the government of Ghana put in place to ensure robust tracking, testing, and isolation/quarantine/treatment of suspected cases in real-time. These actions, including the timely lockdown and scaling up of testing laboratories, have been reported by studies as the catalyst for the reduced disease burden in terms of mortality and severe illness in Ghana [8].

Nonetheless, other factors, such as the inherent characteristics of the cases, could have provided some cushioning and reduced their risk of death and severe illness. Several studies investigating the protagonist nature of intrinsic immunity in infectious disease outbreaks have supported the argument on how inherent individual traits, like nutritional characteristics and innate immunity, are pivotal in reducing disease susceptibility and subsequent severe outcomes [9,10,11,12]. Also, these studies’ findings further indicate that innate immunity could, however, be significantly reduced by ageing [13]. This evidence may substantiate the earlier stated point that intrinsic characteristics of the studied population probably insulated them and reduced their risk of severe illness and deaths.

Moreover, our study population’s mean age (39.9) was relatively lower than most studies that reported higher mortality during the early stages of the COVID-19 outbreak (21, 22), further corroborating the earlier argument on the association between ageing, innate immunity, and disease severity. Nonetheless, our study is mindful of the age and sample size differences between our analysis and the reference studies that could influence the mean age and further oppose our assertion. Also, our hypothesis on ageing and innate immunity may not be absolute as it is not inclusive of other equally important factors, such as the behavioural characteristics of the cases [14,15,16]. Therefore, we suggest that future studies explore how individuals’ lifestyles and behaviours influence the risk of COVID-19 severe illness and deaths.

Furthermore, the findings of our study indicated that increasing age and the presence of diabetes mellitus increase the odds of developing severe COVID-19 disease. These findings corroborate evidence in the literature regarding indicators of severe COVID-19 illness [17,18,19]. Therefore, health systems should continuously provide specific and individual-focused interventions using these findings as a guide to reduce the COVID-19 disease burden.

## Strength and limitations

This article study is one of the few studies to examine the distribution and risk factors for severe COVID-19 disease in Ghana. Consequently, it presents unique findings that could serve as pointers for critical public health interventions in Ghana. For instance, the outcomes of risk factors for severe COVID-19 illness analysis could inform continued prioritized interventions, such as vaccination, to shield high-risk individuals. Concerning limitations, our study included only cases from the main COVID-19 centre; thus, patients in other hospitals scattered around the country were excluded. Also, the study had a relatively smaller sample size. Therefore, the findings may not be generalizable to the broader COVID-19 population in Ghana. Finally, we were limited in drawing causal associations between our explanatory and outcome variables due to the observational nature of our study.

## Conclusion and recommendations

This study aimed to report the epidemiological characteristics of COVID-19 patients admitted at GEMH during the first three months of the viral outbreak in Ghana. Our findings indicated that Ghana’s COVID-19 cases in the early stages of the pandemic were mainly mild to moderate disease and with a comparatively low mortality. This may be due to lessons learnt during the devastating impact of the pandemic in other countries before it reached Ghana our shores. Also, our analysis showed that increasing age and diabetes mellitus are risk factors for severe COVID-19 disease, as reported by several studies in the literature. Therefore, current interventions based on similar evidence are recommended to continue to contain the viral spread. Most importantly, regular research must be encouraged to identify any changes in risk characteristics to proffer updated and appropriate interventions to mitigate the COVID-19 menace holistically.

## Data Availability

The data analysed in this study are available upon request.

## Ethics approval

Ethics approval for this study was given by the Ghana Health Service (GHS) Ethics Review Committee. The ethics approval number is GHS-ERC007/06/20.

## Authors’ contribution

EOM, DA and SC conceptualized the study, SC drafted the first manuscript and conducted the data analysis, SC and DA retrieved the data from GEMH, and EOM and DA reviewed the manuscript. All authors agreed on the final manuscript.

## Acknowledgements

The authors wish to acknowledge the contributions of staff of GEMH, Kwabenya-Ghana.

## Patient and public involvement

No patients or public were directly involved in this study

## Patient consent for publication

Not required.

## Data availability statement

Data is available upon request.

## References

1. World Health Organisation. (2021). Coronacirus. (Online). Available at https://www.who.int/health-topics/coronavirus#tab=tab_1 (Accessed on 29/04/2020).

2. Ghana health service COVID 19 Ghana’s outbreak response management update htttps://ghanahealthservice.org

3. Peipei Wang, Xinqui Zheng, Jiayang Li, Bangren Zhu, prediction of epidemic trends in COVID-19 with logistic model and machine learning technics, chaos, solutions and Fractals, volume 139,20220,110058, ISSN 0960-0779. (https://doi.org/10.1016/j.chaos.2020.110058. (https://www.sciencedirect.com/science/article/pii/s0960077920304550)

4. World Health Organization. (2020). Clinical management of severe acute respiratory infection (SARI) when COVID-19 disease is suspected: Interim guidance. March 2020

5. Yang J, Zheng Y, Gou X, Pu K, Chen Z, Guo Q, Ji R, Wang H, Wang Y, Zhou Y. Prevalence of comorbidities and its effects in patients infected with SARS-CoV-2: a systematic review and meta-analysis. International Journal of Infectious Diseases. 2020 May 1; 94:91–5.

6. Chen N, Zhou M, Dong X, Qu J, Gong F, Han Y, Qiu Y, Wang J, Liu Y, Wei Y, Yu T. Epidemiological and clinical characteristics of 99 cases of 2019 novel coronavirus pneumonia in Wuhan, China: a descriptive study. The lancet. 2020 Feb 15; 395(10223):507–13.

7. Fang L, Karakiulakis G, Roth M. Are patients with hypertension and diabetes mellitus at increased risk for COVID-19 infection? The lancet respiratory medicine. 2020 Apr 1; 8(4): e21.

8. WHO. Report of the WHO-China Joint Mission on Coronavirus Disease 2019 (COVID-19). https://www.who.int/docs/default-source/coronaviruse/who-china-joint-mission-on-covid-19-final-report.pdf. Accessed 18 Mar 2020.

9. Janeway CA Jr, Medzhitov R. Innate immune recognition. Annu Rev Immunol. 2002; 20:197–216. doi: 10.1146/annurev.immunol.20.083001.084359. Epub 2001 Oct 4. PMID: 11861602.

10. Kumar H, Kawai T, Akira S. Pathogen recognition by the innate immune system. Int Rev Immunol. 2011 Feb; 30(1):16–34. doi: 10.3109/08830185.2010.529976. PMID: 21235323.

11. Beutler B. Innate immunity: an overview. Mol Immunol. 2004 Feb; 40(12):845–59. doi: 10.1016/j.molimm.2003.10.005. PMID: 14698223.

12. Dempsey PW, Allison ME, Akkaraju S, Goodnow CC, Fearon DT. C3d of complement as a molecular adjuvant: bridging innate and acquired immunity. Science. 1996 Jan 19; 271(5247):348–50. doi: 10.1126/science.271.5247.348. PMID: 8553069.

13. Opal SM, Girard TD, Ely EW. The immunopathogenesis of sepsis in elderly patients. Clin Infect Dis. 2005 Nov 15;41 Suppl 7: S504–12. doi: 10.1086/432007.PMID:16237654.

14. Inoue Y, Koizumi A, Wada Y, Iso H, Watanabe Y, Date C, Yamamoto A, Kikuchi S, Inaba Y, Toyoshima H, Tamakoshi A. Risk, and protective factors related to mortality from pneumonia among middleaged and elderly community residents: the JACC Study. J Epidemiol. 2007 Nov; 17(6):194–202. doi: 10.2188/jea.17.194. PMID: 18094518; PMCID: PMC7058467.

15. Paulsen J, Askim Å, Mohus RM, Mehl A, Dewan A, Solligård E, Damås JK, Åsvold BO. Associations of obesity and lifestyle with the risk and mortality of bloodstream infection in a general population: a 15-year follow-up of 64 027 individuals in the HUNT Study. Int J Epidemiol. 2017 Oct 1; 46(5):1573–1581. doi: 10.1093/ije/dyx091. PMID: 28637260.

16. Sudlow C, Gallacher J, Allen N, Beral V, Burton P, Danesh J, Downey P, Elliott P, Green J, Landray M, Liu B, Matthews P, Ong G, Pell J, Silman A, Young A, Sprosen T, Peakman T, Collins R. UK biobank: an open access resource for identifying the causes of a wide range of complex diseases of middle and old age. PLoS Med. 2015 Mar 31; 12(3): e1001779. doi: 10.1371/journal.pmed.1001779. PMID: 25826379; PMCID: PMC4380465.

17. Casas-Rojo JM, Antón-Santos JM, Millán-Núñez-Cortés J, Lumbreras-Bermejo C, Ramos-Rincón JM, Roy-Vallejo E, Artero-Mora A, Arnalich-Fernández F, García-Bruñén JM, Vargas-Núñez JA, Freire-Castro SJ, Manzano-Espinosa L, Perales-Fraile I, Crestelo-Viéitez A, Puchades-Gimeno F, Rodilla-Sala E, Solís-Marquínez MN, Bonet-Tur D, Fidalgo-Moreno MP, Fonseca-Aizpuru EM, Carrasco-Sánchez FJ, Rabadán-Pejenaute E, Rubio-Rivas M, Torres-Peña JD, Gómez-Huelgas R; en nombre del Grupo SEMI-COVID-19 Network. Clinical characteristics of patients hospitalized with COVID-19 in Spain: Results from the SEMI-COVID-19 Registry. Rev Clin Esp (Barc). 2020 Nov; 220(8):480–494. doi: 10.1016/j.rce.2020.07.003. Epub 2020 Jul 19. PMID: 32762922; PMCID: PMC7480740.

18. Guan WJ, Ni ZY, Hu Y, Liang WH, Ou CQ, He JX, Liu L, Shan H, Lei CL, Hui DSC, D. B, Li LJ, Zeng G, Yuen KY, Chen RC, Tang CL, Wang T, Chen PY, Xiang J, Li SY, Wang JL, Liang ZJ, Peng YX, Wei L, Liu Y, Hu YH, Peng P, Wang JM, Liu JY, Chen Z, Li G, Zheng ZJ, Qiu SQ, Luo J, Ye CJ, Zhu SY, Zhong NS; China Medical Treatment Expert Group for Covid-19. Clinical Characteristics of Coronavirus Disease 2019 in China. N Engl J Med. 2020 Apr 30; 382(18):1708–1720. doi: 10.1056/NEJMoa2002032. Epub 2020 Feb 28. PMID: 32109013; PMCID: PMC7092819

19. Muniyappa R, Gubbi S. COVID-19 pandemic, coronaviruses, and diabetes mellitus. American Journal of Physiology-Endocrinology and Metabolism. 2020 May 1;318(5): E736–41.

20. Khoo A. Ghana in COVID-19 pandemic. Inter-Asia Cultural Studies. 2020 Oct 1;21(4):542–56.

21. Grasselli G, Greco M, Zanella A, Albano G, Antonelli M, Bellani G, Bonanomi E, Cabrini L, Carlesso E, Castelli G, Cattaneo S. Risk factors associated with mortality among patients with COVID-19 in intensive care units in Lombardy, Italy. JAMA internal medicine. 2020 Oct 1;180(10):1345–55.

22. Di Castelnuovo A, Bonaccio M, Costanzo S, Gialluisi A, Antinori A, Berselli N, Blandi L, Bruno R, Cauda R, Guaraldi G, My I. Common cardiovascular risk factors and in-hospital mortality in 3,894 patients with COVID-19: survival analysis and machine learning-based findings from the multicentre Italian CORIST Study. Nutrition, Metabolism and Cardiovascular Diseases. 2020 Oct 30;30(11):1899–913.

